# Financial burden of Chronic Kidney Disease Patients on Maintenance Hemodialysis in Chittagong, Bangladesh

**DOI:** 10.1101/2021.07.15.21260572

**Authors:** Rajat Sanker Roy Biswas, Jishu Deb Nath, Kazi Farhad Ahmed

## Abstract

**Background:** Chronic kidney disease (CKD) is an important public health problem. Renal replacement therapy (RRT) is needed to patients who goes to end stage renal disease(ESRD). Most of the evidence on its costs relates to patients receiving dialysis or kidney trans-plants, which shows that, in these phases, CKD poses a high burden to payers. The aim of this study was to estimate the financial burden of patients with CKD on maintenance hemodialysis.

**Methods:** It is one -year observational study, carried out to collect data on 105 patients with CKD on dialysis taking from different centers of Chittagong Bangladesh. After collection of sociodemographic informations financial status were collected from patients who gave informed written consent to be included in the study. Data were analyzed by SPSS 20.

**Results:** Regarding gender distribution, male was 73(69.5%) and female was 32(30.5%). Male to female ratio was 2.28:1. Age group distribution revealed 10(9.5%) patients were at <30 years, 18(17.1%) were at 31-40 years, 23(21.9%) were at 41-50 years, 26(24.8%) were at 51-60 years, 20(19.0%) were at 61-70 years and 8(7.6%) were >71 years age. Among all most were involved in service 40(38.1%) and business 23(21.9%). Socio economic status of the patients revealed 42(40.0%) patients were from upper middle class, 50(47.6%) were from lower middle class. Among all, 33(31.4%) patients took treatment from abroad. Regarding bearing of cost of the dialysis 24(22.9%) were self financed and 35(33.3%) got help from others(non family member), 11(10.5%) took loan and 13(12.4%) sold their stable land property. Regarding tenure of dialysis 31(29.5% patients are getting dialysis <1 year and 74(70.5%) were getting it for 1-2 years. Previous monthly income before start of dialysis was zero(0) in 33(31.4%) patients and it was >30000 taka/month in 34(32.4%) patients and after start of dialysis present monthly income was zero(o) per month in 67(63.8%) patients and >30000 taka/month in 13(12.4%) patients. Regarding expenditure for each dialysis showed 2(1.9%) had zero(0) taka and 46(43.8%) patients needed 1000-2000 taka/session and 41(39.0%) needed 2000-3000 taka /session.

**Conclusion:** Dialysis in CKD patients is a huge financial burden to the patients himself and the family. Government assistance should be provided to all patients who are unable to bear the cost.

## Introduction

Chronic kidney disease(CKD) is a major global public health issue, affecting over 10% of the population worldwide^1^. The problem was ranked 16^th^ among the leading causes of death in 2016, and is expected to rise to 5^th^ ranked by 2040^2^. Chronic kidney disease is defined as an abnormality of kidney function or structure for 3 months^3^ and is a significant burden for individuals, health care systems and societies; it is associated with increased hospitalization, productivity loss, morbidity and early mortality.

Renal replacement therapy (RRT), through either dialysis or renal transplantation, is a lifesaving. But it is a high-cost treatment for people with end-stage kidney disease. It has been available in high-income countries for more than 50 years, but a lower middle income country like Bangladesh RRT is still unavailable for all patients who need it. The use of dialysis to treat end-stage kidney disease varies substantially between regions, probably because of differences in population demographics, prevalence of end-stage kidney disease, and factors affecting access to and provision of RRT.^1^

Lots of patients cannot afford the RRT due to high cost in relation to their income. Some get financial helps from social charity but it is not enough to maintain regular dialysis. Government assistance also are not up to the mark. So the objective of this present study was to observe the financial burden of CKD patients who are on maintenance hemo dialysis in Chittagong, Bangladesh.

## Methods

Present observational study was done among ESRD patients who take hemodialysis in different centers of Chittagong, Bangladesh. Patients who were enlisted in the dialysis center and took at least dialysis for one month were enrolled in the study after informed written consent. A total of 105 patients were included in the study from three dialysis center from Chittagong. After enrollment history was taken regarding socio demographic information and later financial status were collected from patients or from their legal guardian if patient was very sick. Financial data were collected using Bangladeshi Taka as a monetary unit Later after collection of all data it was analyzed by SPSS 20.

## Results

Regarding gender distribution of study patients male was 73(69.5%) and female was 32(30.5%). Male to female ratio was 2.28:1.(Table 1) Age group distribution revealed 10(9.5%) patients were at <30 years, 18(17.1%) were at 31-40 years, 23(21.9%) were at 41-50 years, 26(24.8%) were at 51-60 years, 20(19.0%) were at 61-70 years and 8(7.6%) were >71 years age.(Table 2)Among all most were involved in service 40(38.1%) and business 23(21.9%) and among female most were house wives 29(27.6%).(Table 3) Socio demographic status of the patients revealed 42(40.0%) patients were from upper middle class. 50(47.6%) were from lower middle class and 12(11.4%) were poor. Only one patient was found as rich.(Table 4) Among all 33(31.4%) patients took treatment from abroad. (Table 5). Regarding bearing cost 24(22.9%) were self financed and 35(33.3%) got help from other(non family member), 11(10.5%) took loan and 13(12.4%) sold their stable land property(Table 6) Regarding tenure of dialysis 31(29.5% patients are getting dialysis <1 year and 74(70.5%) were getting it for 1-2 years.(Table 7) Regarding previous monthly income before start of dialysis was zero(0) in 33(31.4%) patients and it was >30000 taka/month in 34(32.4%) patients and fter start of dialysis present monthly income was zero(o) per month in 67(63.8%) patients and >30000 taka/month in 13(12.4%) patients(Table 8 and table 9).Regarding expenditure for each dialysis on 2(1.9%) had zero(0) taka and 46(43.8%) patients needed 1000-2000 taka/session and 41(39.0%) needed 2000-3000 taka /session.(Table 10)

**Table 1:** Gender distribution

|  | Frequency | Percent |
| --- | --- | --- |
| Male | 73 | 69.5 |
| Female | 32 | 30.5 |
| Total | 105 | 100.0 |

**Table 2:** Age group distribution

|  | Frequency | Percent |
| --- | --- | --- |
| <30 years | 10 | 9.5 |
| 31-40 years | 18 | 17.1 |
| 41-50 years | 23 | 21.9 |
| 51-60 years | 26 | 24.8 |
| 61-70 years | 20 | 19.0 |
| >71 years | 8 | 7.6 |
| Total | 105 | 100.0 |

**Table 3:** Occupation of the patients before dialysis

|  | Frequenc<br>y | Percent |
| --- | --- | --- |
| Service | 40 | 38.1 |
| Student | 3 | 2.9 |
| House wife | 29 | 27.6 |
| Day labor | 1 | 1.0 |
| Business | 23 | 21.9 |
| Retired person | 9 | 8.6 |
| Total | 105 | 100.0 |

**Table 4:** Socioeconomic status

|  | Frequenc<br>y | Percent |
| --- | --- | --- |
| Rich | 1 | 1.0 |
| Upper middle<br>class | 42 | 40.0 |
| Lower middle<br>class | 50 | 47.6 |
| Poor | 12 | 11.4 |
| Total | 105 | 100.0 |

**Table 5:** Treatment taken from abroad

|  | Frequency | Percent |
| --- | --- | --- |
| Yes | 33 | 31.4 |
| No | 72 | 68.6 |
| Total | 105 | 100.0 |

**Table 6:** Cost of treatment bearded

|  | Frequency | Percent |
| --- | --- | --- |
| Father | 6 | 5.7 |
| Help from others(Non family member) | 35 | 33.3 |
| Husband | 11 | 10.5 |
| Loan | 11 | 10.5 |
| Self | 24 | 22.9 |
| Selling of stable land property | 13 | 12.4 |
| Social fund | 4 | 3.8 |
| Son | 1 | 1.0 |
| Total | 105 | 100.0 |

**Table 7:** Tenure of dialysis grading

|  | Frequency | Percent |
| --- | --- | --- |
| < 1 year | 31 | 29.5 |
| 1-2 years | 74 | 70.5 |
| Total | 105 | 100.0 |

**Table 8:** Previous monthly income before dialysis started

|  | Frequency | Percent |
| --- | --- | --- |
| 0 | 33 | 31.4 |
| 1-10000 taka/month | 9 | 8.6 |
| 10000-20000 taka/month | 18 | 17.1 |
| 20000-30000 taka/month | 11 | 10.5 |
| >30000 taka/month | 34 | 32.4 |
| Total | 105 | 100.0 |

1 USD= 85 taka

**Table 9:** Present monthly income after dialysis started

|  | Frequenc<br>y | Percent |
| --- | --- | --- |
| 0 | 67 | 63.8 |
| 1-10000 taka/month | 8 | 7.6 |
| 10000-20000<br>taka/month | 9 | 8.6 |
| 20000-30000<br>taka/month | 8 | 7.6 |
| >30000 taka/month | 13 | 12.4 |
| Total | 105 | 100.0 |

1 USD= 85 taka

**Table 10:** Expenditure for each dialysis.

|  | Frequenc<br>y | Percent |
| --- | --- | --- |
| 0 taka | 2 | 1.9 |
| 1-1000 taka/session | 16 | 15.2 |
| 1000-2000<br>taka/session | 46 | 43.8 |
| 2000-3000<br>taka/session | 41 | 39.0 |
| Total | 105 | 100.0 |

1 USD= 85 taka

## Discussion

Regarding gender distribution of study patients male was 73(69.5%) and female was 32(30.5%). Male to female ratio was 2.28:1. Bangladesh is a country of low educational level and male gets more preference than female. Hence, here, more males come under dialysis than female. Age group distribution revealed most were distributed in middle age group. A previous study^4^ showed that the majority of patients in their study were at age group50–60years,

We found most were involved in service 40(38.1%) and business 23(21.9%). Socio demographic status of the patients revealed 42(40.0%) patients were from upper middle class. 50(47.6%) were from lower middle class. Studies.^5,6^ done in India analyzed their demographic data, socioeconomic profile, and etiologies of CKD from different zones. The overall age was 50.1±14.6years, and 36,745(70.3%) of the subjects were males. Overall, females with CKD were 2½ years younger than males(50.9±14.6vs. 48.3±14.4years).^5^ Comparison of data from different zones showed small but statistically significant differences in age distribution and sex ratio. As here, Indian data are near similar to Bangladesh in terms of demographic variables.

Among all 33(31.4%) patients took treatment from abroad. Patients used to visit neibouring country for second opinion and also for better health care in case of different chronic diseases. His is true for the CKD patents also. It causes a big financial impact among the patients in terms of visa processing and transport.

Regarding bearing cost 24(22.9%) were self financed and 35(33.3%) got help from other(non family member), 11(10.5%) took loan and 13(12.4%) sold their stable land property. The risk of chronic kidney disease is bi-directionally affected by level of economic development. Poverty increases the risk of disorders that predispose chronic kidney disease to develop or progress, and worsens outcomes in those who already have chronic kidney disease. Prosperity increases access to RRT. Maintaining health with CKD by dialysis is a challenge for a patient from the third world country like Bangladesh. An analysis of National Health and Nutrition Examination Survey data showed that poverty is associated with an increased risk of proteinuria even after correction for age, sex, ethnic origin, education, obesity, hypertension, diabetes, decreased glomerular filtration rate, and medication use.^7^ People in the lowest socioeconomic quartile are at a 60% greater risk of progressive chronic kidney disease than are those who are in the highest quartile.^8^

Regarding tenure of dialysis 31(29.5% patients are getting dialysis <1 year and 74 (70.5%) were getting it for 1-2 years. Regarding expenditure for each dialysis showed 2(1.9%) had zero(0) taka and 46(43.8%) patients needed 1000-2000 taka/session and 41(39.0%) needed 2000-3000 taka /session

When the duration and tenure of dialysis increases financial burden also increases to the family members and self.

Previous monthly income before start of dialysis was zero (0) in 33(31.4%) patients and it was >30000 taka/month in 34(32.4%) patients and after start of dialysis present monthly income was zero (0) per month in 67(63.8%) patients and >30000 taka/month in 13(12.4%) patients. It is clearly seen that previous income before dialysis was more than the income after start of dialysis. Every time the patient undergoes dialysis it sucks the patient and gradually make the patient poor.

So, in a country like Bangladesh, CKD patients should get government assistance for regular dialysis. There should be a national guideline for helping the CKD patients on MHD. A national database should be made, adequate RRT center should made and by that only most of the patients can get dialysis and can live a anxious free life.

## Data Availability

Data is available if asked

